# Blood-Based Brain Injury Biomarkers to Prognosticate Outcome after Pediatric Cardiac Arrest

**DOI:** 10.1101/2022.02.28.22271632

**Authors:** Ericka L Fink, Patrick M. Kochanek, Ashok Panigrahy, Sue R. Beers, Rachel P. Berger, Hülya Bayir, Jose Pineda, Christopher Newth, Alexis A Topjian, Craig A. Press, Aline B. Maddux, Frederick Willyerd, Elizabeth A Hunt, Jordan Duval-Arnould, Ashley Siems, Melissa G Chung, Lincoln Smith, Jesse Wenger, Leslie Doughty, J. Wesley Diddle, Jason Patregnani, Juan Piantino, Karen Hallermeier Walson, Binod Balakrishnan, Michael T. Meyer, Stuart Friess, David Maloney, Pamela Rubin, Tamara L. Haller, Amery Treble-Barna, Chunyan Wang, Robert R.S.B. Clark, Anthony Fabio, the POCCA Investigators

## Abstract

**Background:** Prognostication after cardiac arrest in children is challenging due to a lack of validated methods to evaluate direct brain injury. The objective of this multicenter study was to analyze biomarker accuracy to prognosticate outcome 1 year post-arrest.

**Methods:** Fourteen U.S. centers enrolled 164 children ages 48 h - 17 years with pre-arrest Pediatric Cerebral Performance Category score of 1-3 who were admitted to an intensive care unit after cardiac arrest. Glial fibrillary acidic protein (GFAP), ubiquitin carboxyl-terminal esterase-L1 (UCH-L1), neurofilament light (NfL), and Tau concentrations were measured in blood samples from post-arrest days 1-3 using Quanterix™ Simoa 4-Plex assay, Billerica, MA. Unfavorable outcome was death or survival with Vineland Adaptive Behavioral Scale-Third Edition score < 70 at 1 year. We analyzed area under receiver operator curve (AUROC) and performed multivariate logistic regressions to determine the association of each biomarker with outcome on days 1-3.

**Results:** Fifty of 120 children with primary outcomes available had an unfavorable outcome, including 43 deaths. Compared to those with favorable outcomes, more children with unfavorable outcome had out-of-hospital (36% vs. 70%) and unwitnessed (7% vs. 46%) events, p<0.05. For days 1-3, concentrations of all four measured biomarkers were increased in children with an unfavorable vs. favorable outcome, p<0.05. On post-arrest day 1, NfL demonstrated the best outcome classification (AUROC 0.731 [95% confidence interval 0.642, 0.820]) while UCH-L1 performed best on days 2 (0.860 [0.785, 0.935]) and 3 (0.837 [0.747, 0.926]). After covariate adjustment, NfL concentrations on day 1 (odds ratio 5.9 [95% confidence interval 1.8, 19.2], day 2 (11.9 [3.8, 36.9]), and day 3 (10.2 [3.1, 33.3]), UCH-L1 on day 2 (11.3 [3.0, 42.4]) and day 3 (7.6 [2.1, 27.1]), GFAP on day 2 (2.3 [1.2, 4.5]) and day 3 (2.2 [1.2, 4.0]), and tau on day 1 (2.4 [1.1, 5.3]), day 2 (2.3 [1.3, 4.0]), and day 3 (2.0 [1.2, 3.6]) were associated with unfavorable outcome, p<0.05.

**Conclusions:** Blood-based brain injury biomarkers accurately prognosticated death or unfavorable adaptive behavior composite outcome at 1 year after pediatric cardiac arrest. Accuracy of biomarkers to predict neurodevelopmental outcomes beyond 1 year should be evaluated.

**Clinical Trial Registration:** URL: https://www.clinicaltrials.gov: Unique identifier: NCT02861534

**Clinical Perspective:** *What is new?:*

- In children who suffered a cardiac arrest, post-arrest blood levels of neurofilament light, ubiquitin carboxyl-terminal esterase-L1, glial fibrillary acidic protein, and tau predicted death or unfavorable adaptive behavior composite outcome at 1 year.
- Neurofilament light was the best performing biomarker to predict outcome on day 1 while ubiquitin carboxyl-terminal esterase-L1 performed best on days 1 and 2.

*What are the clinical implications?:*

- Blood-based brain injury biomarkers should be considered for clinical use to aid in prognostication after pediatric cardiac arrest.
- Biomarker levels should be assessed as tools to aid in the prediction of neurodevelopmental outcomes beyond one year.

## Introduction

Approximately 10,000 children experience in-or out-of-hospital cardiac arrest annually in the United States (US)^1, 2^. Children with return of spontaneous circulation are at high risk of neurologic morbidity and death due to global hypoxic-ischemic brain injury^3^. Early prognostication could provide critical information regarding a child’s neurologic status to support clinician and family decision-making and treatment. However, a standardized approach to prognosticate neurologic outcomes with accuracy early in the hospital course is lacking^4^.

Small observational studies demonstrate the potential for blood-based brain injury biomarkers to predict outcome after pediatric cardiac arrest^5, 6^. Brain-specific biomarkers ubiquitin carboxyl-terminal esterase L1 (UCH-L1) and glial fibrillary acidic protein (GFAP) are approved by the U.S. Federal and Drug Administration to assist in clinical decision-making in mild traumatic brain injury in adults and differentiated outcomes in a pilot study in pediatric cardiac arrest^7-9^. Under normal conditions, UCH-L1, located in neurons, participates in the degradation of damaged proteins^10^ and GFAP forms intermediate filaments that support shape and function of astroglia cells^11^. Furthermore, it was shown that blood-based white matter injury biomarkers may serve as prognostic tools after pediatric and adult cardiac arrest^12, 13^. Thus, neurofilament light (NfL), a component of the neuronal cytoskeleton providing structural support to axons^14-16^ and tau, a microtubule-stabilizing neuro-axial protein that maintains stability of axon microtubules, were also included in this study^17, 18^.

We evaluated the association and accuracy of these four brain-specific biomarkers on days 1-3 post-pediatric cardiac arrest to predict death or unfavorable adaptive behavior composite outcome at 1 year in a prospective multicenter, observational study. We hypothesized that each blood-based biomarker could accurately predict outcome after cardiac arrest.

## Methods

Data that support the findings of this study are available from the corresponding author upon reasonable request.

### Regulatory

The University of Pittsburgh Institutional Review Board (IRB) served as the study’s central IRB and approved the study (STUDY20110318) at the UPMC Children’s Hospital of Pittsburgh. Two sites, Children’s Healthcare of Atlanta (IRB #16-204) and Children’s Wisconsin (IRB #2 FWA00001809) obtained independent IRB approval due to inability to participate in the central IRB.

### Study design and setting

This prospective, observational study was performed at 14 academic, referral center pediatric intensive care units (PICUs) in the US. Enrollment was conducted between May 2017 and August 2019, with follow-up through one-year post-enrollment.

### Participants

Eligible children were between the ages of 48 hours -17 years, had chest compressions performed for any duration of time for in-or out-of-hospital events, and had a pre-cardiac arrest Pediatric Cerebral Performance Category score of 1-3^19^. Children were excluded if they had a do not resuscitate order; were in foster care or the judicial system; were pregnant; were actively undergoing brain death evaluation; or if the staff were unable to obtain a blood sample (200 uL) within 24 hours of the arrest. Study coordinators and investigators screened for eligible patients daily in the PICU and via the electronic medical record. Some centers received automated or custom alerts from clinicians when a cardiac arrest patient was admitted. Informed consent was obtained from the patient’s parent or guardian and patient assent was obtained, when appropriate based on age and clinical status. Patients did not undergo any study-related treatment interventions. Clinical care was provided per the patient’s clinical team.

### Data Collection

Data were collected locally guided by the Case Report Form (CRF). The Utstein template for cardiac arrest was used to define cardiac arrest related variables^20^. Patient demographics (e.g., age, sex, race), cardiac arrest and resuscitation (e.g., etiology and location of cardiac arrest, number of epinephrine doses), and post-resuscitation care and test results (e.g., mechanical ventilation, vasoactive support, initial blood lactate), and outcomes (e.g., hospital disposition) were collected. Cause of death was abstracted from the death certificate.

### Blood-based biomarkers

Three milliliters of blood were collected prospectively on days 1, 2, and 3 after return of spontaneous circulation. Day 1 was defined as within the first 24 hours post-return of spontaneous circulation; day 2 was 24-48 hours, and day 3 was 48-72 hours. Blood samples were centrifuged, aliquoted, frozen at -70°C, and mailed to the University of Pittsburgh for storage and batched analysis. If prospective samples were not possible, leftover serum or plasma could be obtained from the hospital’s laboratory and stored and mailed as above. NfL, UCH-L1, GFAP, and Tau concentrations in serum or plasma were measured by a laboratory technician at Quanterix (Billerica, MA) blinded to patient data using the Simoa® Human Neurology 4-plex assay. Lower limits of quantitation from Quanterix were 0.241 pg/ml (NfL), 5.45 pg/ml (UCH-L1), 0.467 pg/ml (GFAP), and 0.053 pg/ml (Tau). Samples were measured in duplicate; average concentrations were used for analysis. Dilution factors of 4, 10, or 1,000 were used. Clinical team members were blinded to the biomarker results.

### Outcome Measures

Patient follow-up was performed by each center. Parents or guardians completed the Vineland Adaptive Behavior Scale, Third Edition (VABS) rating form at 1 year via in-person, mail, or phone follow-up^21^. The VABS is a standardized measure of adaptive behavior function based on caregiver report of daily functioning. Study coordinators reviewed the patient’s medical chart to determine 1-year survival prior to contacting families. The VABS provides age-corrected standard scores (mean [SD], 100 [15]) for individuals from birth through 90 years in 4 domains (communication, daily living, socialization, and motor skills) and an overall adaptive behavior composite, with higher scores denoting better functioning. Favorable outcome was defined as survival with a VABS overall adaptive behavior composite of ≥ 70 and unfavorable outcome was defined as a VABS overall adaptive behavior composite <70 or death, consistent with prior studies of children with cardiac arrest^22, 23^. Questionnaire responses were evaluated for quality and reliability by the primary study team.

### Statistical Analyses

We attest that the first and senior authors had full access to all the data in the study and take responsibility for its integrity and the data analysis. Frequencies and percentages were reported for categorical variables. Biomarker data were log-transformed and are presented as medians (interquartile range [IQR]) due to non-parametric distributions. Differences in categorical data were assessed using chi-square analysis with Fisher’s Exact test, as appropriate. All p-values were 2-sided, and a p<0.05 was considered statistically significant. Area under receiver operator curves (AUROC) and sensitivity/specificity analyses were performed for each biomarker on days 1-3 for outcome. Favorable and unfavorable outcome group differences were compared using the Wilcoxon Rank Sum and Kruskal-Wallis tests. Biomarker specificity, threshold level (pg/ml), and sensitivity on univariate biomarker models were performed with set specificity of 95% to minimize the chance of false prognostication of unfavorable outcome.

Multivariate logistic regression models with stepwise selection with entry and stay level of 0.20 and forced inclusion of log biomarker concentrations were used to evaluate the association between individual biomarker concentrations on each day and one-year outcome. All patient and cardiac arrest variables evaluated for inclusion in multivariate analysis. Missing data were not imputed. All analyses were conducted using SAS® 9.2 (Copyright © [2002-2008] SAS Institute Inc., Cary, NC, USA).

## Results

### Participants

Of 932 children who had a cardiac arrest, 353 were eligible and 164 families consented (Figure 1). One child’s family withdrew all data from the study. One child did not have blood available on day 1 leaving 162 samples available for analysis. On days 2 and 3, samples were available from 141 and 128 patients, respectively. Of 120 children with the primary outcome available at 1 year, 79 (66%) survived. Of the 43 children that died, 2 died after hospital discharge and before the 1 year outcome time point. Of the survivors, 70 (89%) had favorable and 9 (11%) had unfavorable outcome. Thus, overall, 70 (58%) patients had a favorable outcome and 50 (42%) had an unfavorable outcome.

**Figure 1.**
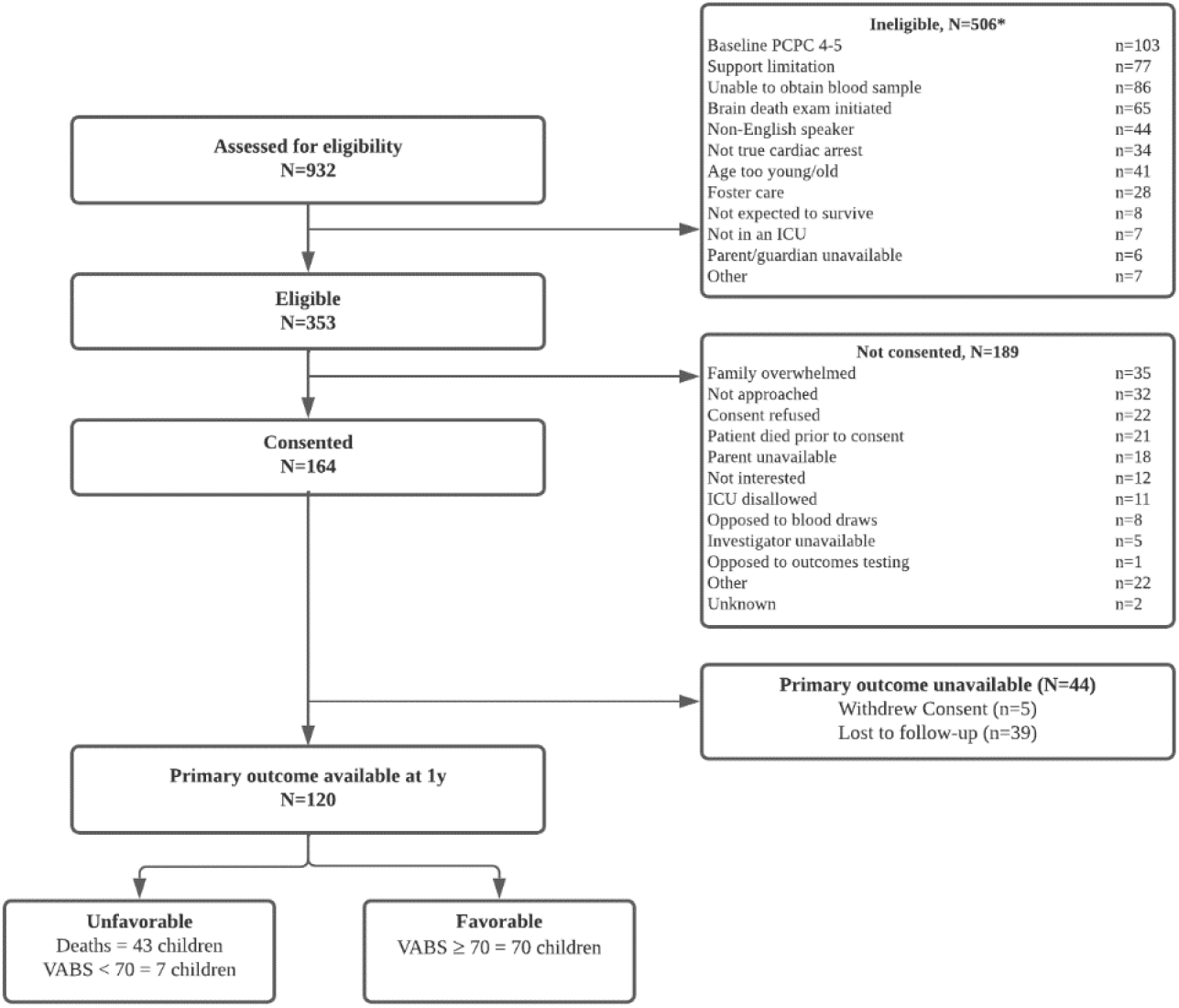
Study flowchart. ^*^Patients may have one or more criteria that makes them ineligible PCPC, Pediatric Cerebral Performance Category; ICU, intensive care unit; VABS, Vineland Adaptive Behavioral Scale, 3^rd^ edition

Overall, the median (IQR) age was 1.0 (0.0, 8.5) years and 41% were female. Patient characteristics were not different in children with favorable or unfavorable outcome (Table 1). Sixty-eight percent of children had a preexisting condition, the most common being cardiovascular (22%). Children with an unfavorable outcome most frequently had out-of-hospital cardiac arrest (70% vs. 36%), longer cardiopulmonary resuscitation (CPR) duration (20 [6, 40] vs. 5 [2, 11] minutes), and an unwitnessed event (46% vs. 7%) than children with favorable outcome (Table 2).

**Table 1.** Patient characteristics overall and by favorable and unfavorable outcome groups.

| n (%) or Median (interquartile range) | Overall<br>N=120 | Favorable<br>N=70 | Unfavorable<br>N=50 | p-value |
| --- | --- | --- | --- | --- |
| **Age, years** | 1.0 (0.0, 8.5) | 1.0 (0.0, 9.0) | 1.0 (0.0, 6.0) | 0.550 |
| **Female sex** | 49 (40.8) | 28 (40.0) | 21 (42.0) | 0.826 |
| **Race** |  |  |  | 0.710 |
| White | 81 (67.5) | 46 (65.7) | 35 (70.0) |  |
| Black | 19 (15.8) | 12 (17.1) | 7 (14.0) |  |
| Asian | 5 (4.2) | 2 (2.9) | 3 (6.0) |  |
| Unknown | 15 (12.5) | 10 (14.3) | 5 (10.0) |  |
| **Hispanic Ethnicity** | 11 (10.0), n=110 | 6 (9.5), n=63 | 5 (10.6), n=47 | 0.847 |
| **Pre-existing conditions<sup>1</sup>** | 77 (68.1) | 44 (69.8) | 33 (66.0) | 0.366 |
| Cardiovascular | 25 (22.1) | 16 (25.4) | 9 (18.0) |  |
| Congenital | 14 (12.4) | 8 (12.7) | 6 (12.0) |  |
| Premature birth | 8 (7.1) | 6 (9.5) | 2 (4.0) |  |
| Neurologic | 7 (6.2) | 3 (4.8) | 4 (8.0) |  |
| Pulmonary | 6 (5.3) | 5 (7.9) | 1 (2.0) |  |
| Oncologic | 3 (2.7) | 2 (3.2) | 1 (2.0) |  |
| Organ or cellular transplant | 3 (2.7) | 1 (1.6) | 2 (4.0) |  |
| Other | 11 (9.7) | 3 (4.8) | 8 (16.0) |  |

Note: Values expressed as n (%), mean ± standard deviation and/or median (25<sup>th</sup>, 75<sup>th</sup> percentiles)
p-value are based on Chi-square test for categorical variables, Kruskal-Wallis test for continuous variables.
Favorable outcome was Vineland Adaptive Behavioral Scale-III (VABS) score $\geq 70$ and Unfavorable outcome was VABS < 70 or died
<sup>1</sup> Patients may have more than one pre-existing condition

**Table 2.** Cardiac arrest, resuscitation, post-arrest, and outcome details overall and by favorable and unfavorable outcome groups.

| n (%) or Median (interquartile range) | **Overall**<br>**N=120** | **Favorable**<br>**N=70** | **Unfavorable**<br>**N=50** | **p-**<br>**value** |
| --- | --- | --- | --- | --- |
| **Primary etiology** | N=107 | N=66 | N=41 | 0.781 |
| Asphyxia | 74 (69.2) | 45 (68.2) | 29 (70.7) |  |
| Cardiac | 33 (30.8) | 21 (31.8) | 12 (29.3) |  |
| **Location, out-of-hospital** | 60 (50.0) | 25 (35.7) | 35 (70.0) | <0.001 |
| **Duration of cardiopulmonary resuscitation, min** | 7.0 (3.0, 20.0),<br>n=99 | 5.0 (2.0, 11.0), n=64 | 20.0 (6.0, 40.0),<br>n=35 | <0.001 |
| **Total number of epinephrine doses** | 0.0 (0.0, 1.0),<br>n=105 | 0.0 (0.0, 0.0), n=63 | 0.5 (0.0, 2.0), n=42 | <0.001 |
| **Defibrillated** | 13 (12.9), n=101 | 8 (13.1), n=61 | 5 (12.5), n=40 | 0.928 |
| **First monitored rhythm** | N=96 | N=58 | N=38 | 0.042 |
| Sinus bradycardia | 33 (34.4) | 23 (39.7) | 10 (26.3) |  |
| Pulseless electrical activity | 23 (24.0) | 14 (24.1) | 9 (23.7) |  |
| Asystole | 19 (19.8) | 7 (12.1) | 12 (31.6) |  |
| Ventricular tachycardia or fibrillation | 15 (15.6) | 10 (17.2) | 5 (13.2) |  |
| Other<sup>1</sup> | 6 (6.3) | 4 (6.9) | 2 (5.3) |  |
| **Witnessed** | 92 (76.7) | 65 (92.9) | 27 (54.0) | <0.001 |
| **Bystander resuscitation** | 42 (35.0) | 17 (24.3) | 25 (50.0) | 0.004 |
| **Hospital length of stay, d** | 18.00 (6.50, 35.50) | 20.50 (10.00, 41.00) | 12.00 (5.00, 34.00) | 0.092 |
| **Intensive care unit length of stay, d** | 12.00 (5.00, 25.00)<br>N=117 | 14.00 (6.00, 21.00)<br>N=69 | 11.00 (5.00, 31.50)<br>N=48 | 0.779 |
| **Disposition at hospital discharge** |  |  |  | <0.001 |
| Home | 55 (45.83) | 50 (71.43) | 5 (10.00) |  |
| Died | 41 (34.17) | 0 (0) | 41 (82.00) |  |
| Inpatient rehabilitation | 17 (14.17) | 15 (21.43) | 2 (4.00) |  |
| Transfer to other hospital | 2 (1.67) | 2 (2.86) | 0 (0) |  |
| Long term care facility | 5 (4.17) | 3 (4.29) | 2 (4.00) |  |
| **Days from cardiac arrest event to death** | 10 (3, 25)<br>N=43 | NA | 10 (3, 25)<br>N=43 |  |
| **Cause of death** | N=43 | NA | N=43 |  |
| Multiple Organ Failure | 13 (30.23) | NA | 13 (30.23) |  |
| Brain death | 11 (25.58) | NA | 11 (25.58) |  |
| Neurologic injury | 11 (25.58) | NA | 11 (25.58) |  |
| Cardiovascular | 8 (18.60) | NA | 8 (18.60) |  |

p-value are based on Chi-square test for categorical variables, Kruskal-Wallis test for median
Favorable outcome was Vineland Adaptive Behavioral Scale-III (VABS) score $\geq 70$ and Unfavorable outcome was VABS $< 70$ or died
<sup>1</sup> Normal sinus, sinus tachycardia, junctional ectopic tachycardia; <sup>2</sup> Per death certificate

### Post-resuscitation data and outcomes

Nearly all (98%) children were mechanically ventilated and vasoactive infusions were frequently used for post-resuscitation support (Supplemental table 1). Extracorporeal membrane oxygenation (ECMO) was used in 23% of children, with extracorporeal-CPR (eCPR) representing the majority of indications (79%).

Targeted temperature management (TTM) was used to prevent fever more frequently in the unfavorable outcome group (36% vs. 17%, p=0.032) while TTM for therapeutic hypothermia was applied in 17% of favorable vs. 6% of unfavorable outcome group patients, p=0.069.

Among non-survivors, 26% of children met criteria for brain death and another 26% had severe neurologic injury.

### Blood biomarker concentrations by outcome group

Biomarker concentrations in pg/ml on days 1, 2, and 3 post-cardiac arrest by outcome group are provided in Supplemental Table 2 (a-c) and as log-transformed concentrations in Figure 2 (a-d). Biomarker concentrations were increased in children with unfavorable vs. favorable one-year outcome for each of the 4 biomarkers studied, p<0.001.

**Figure 2.**
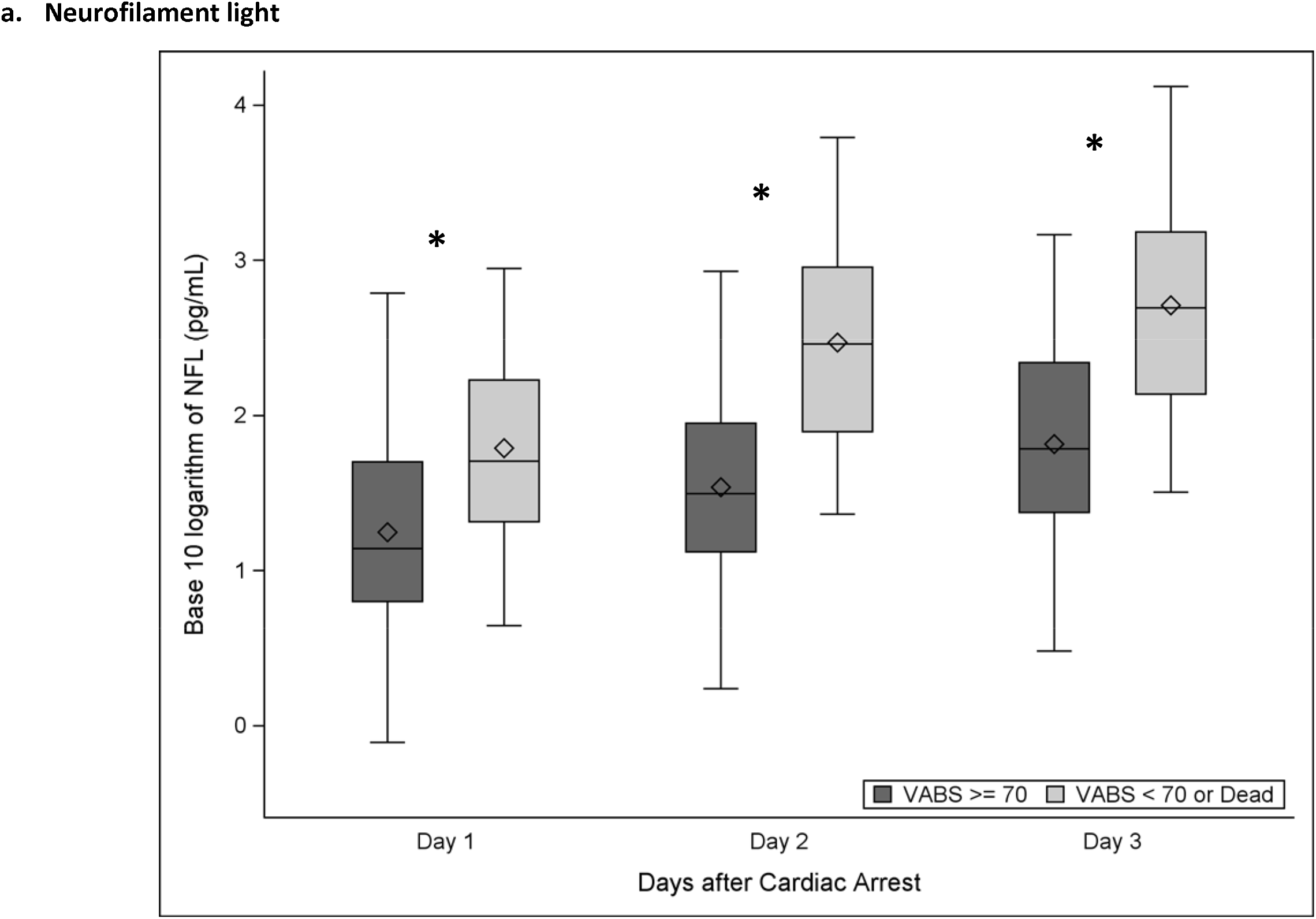
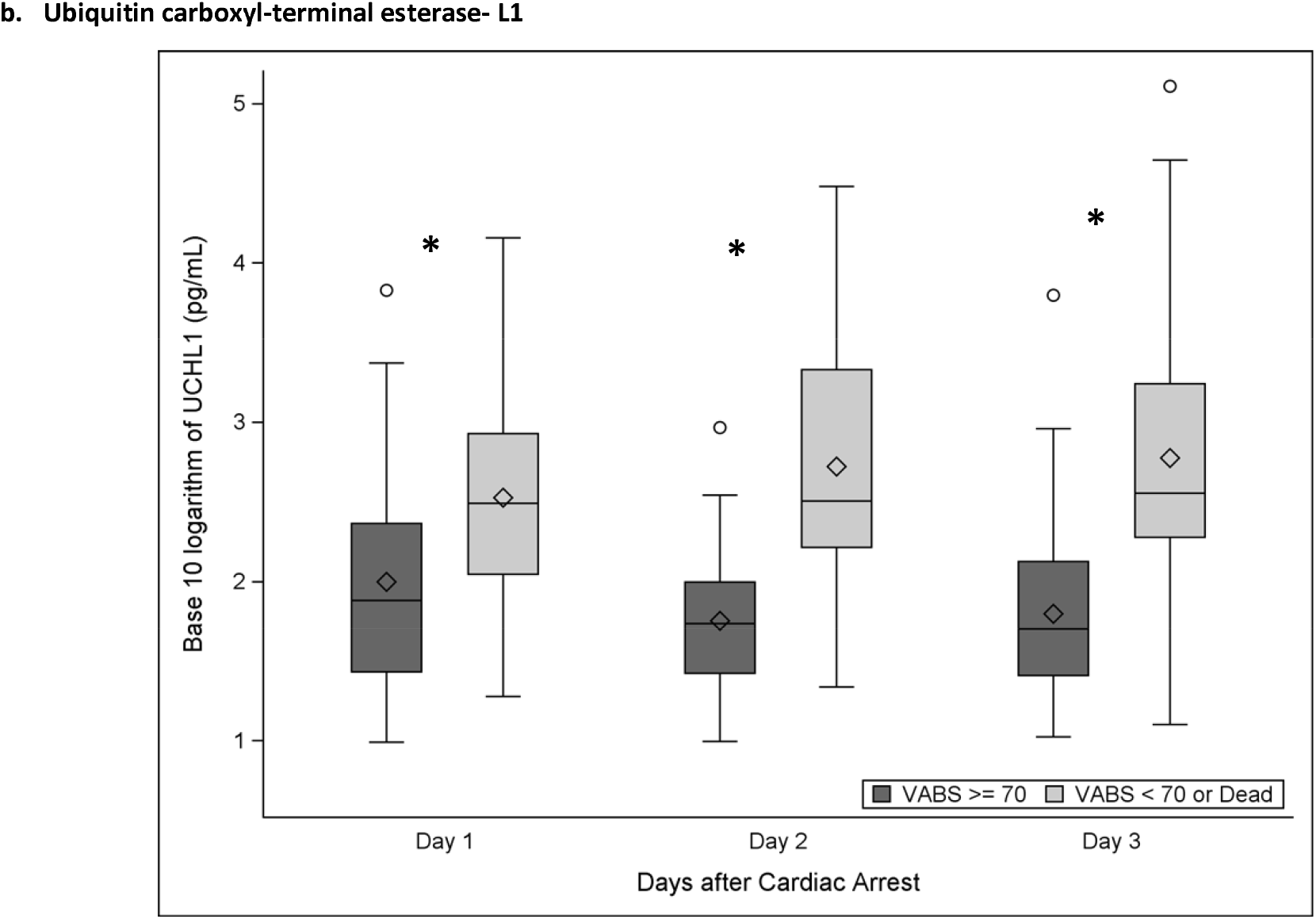
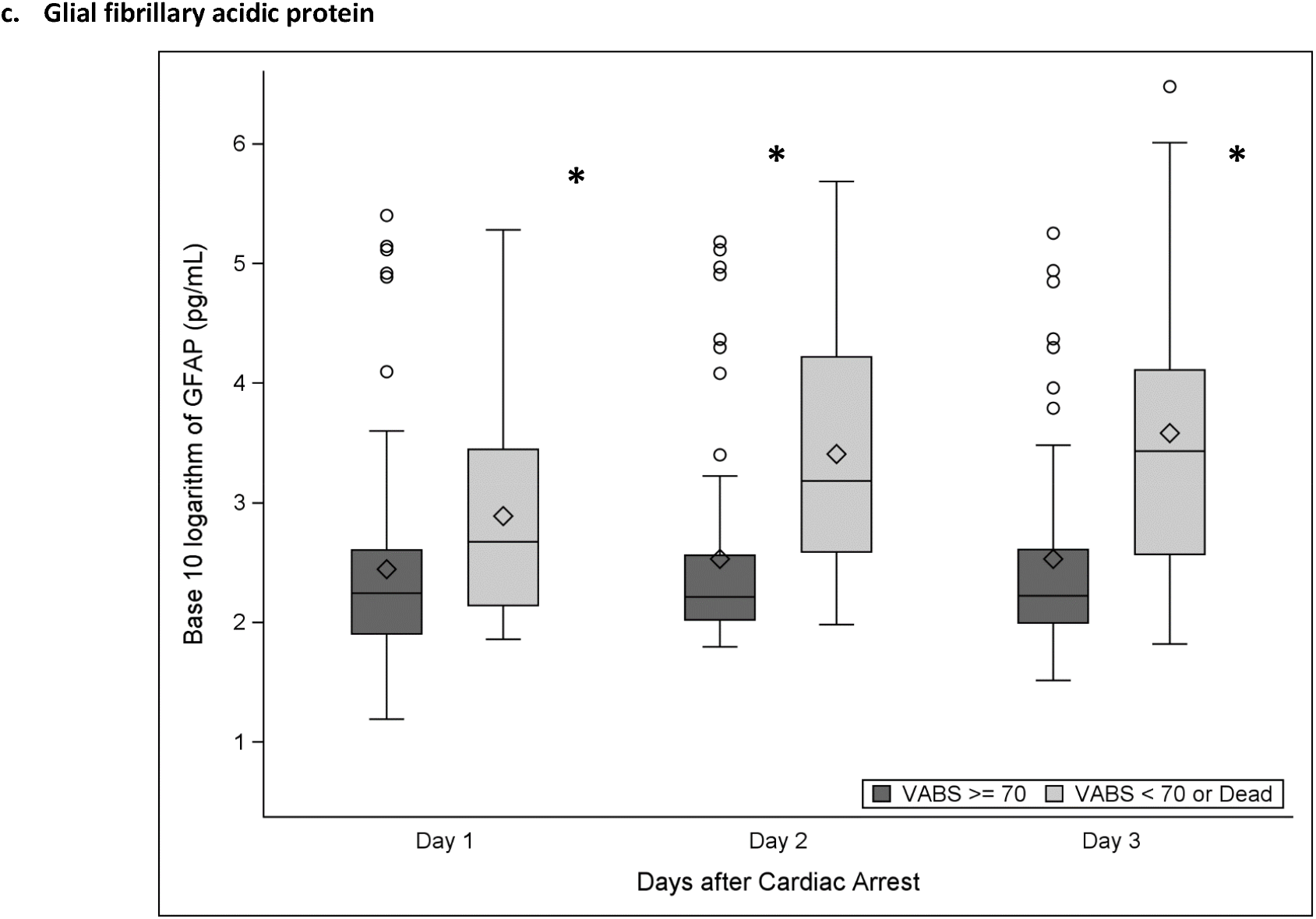
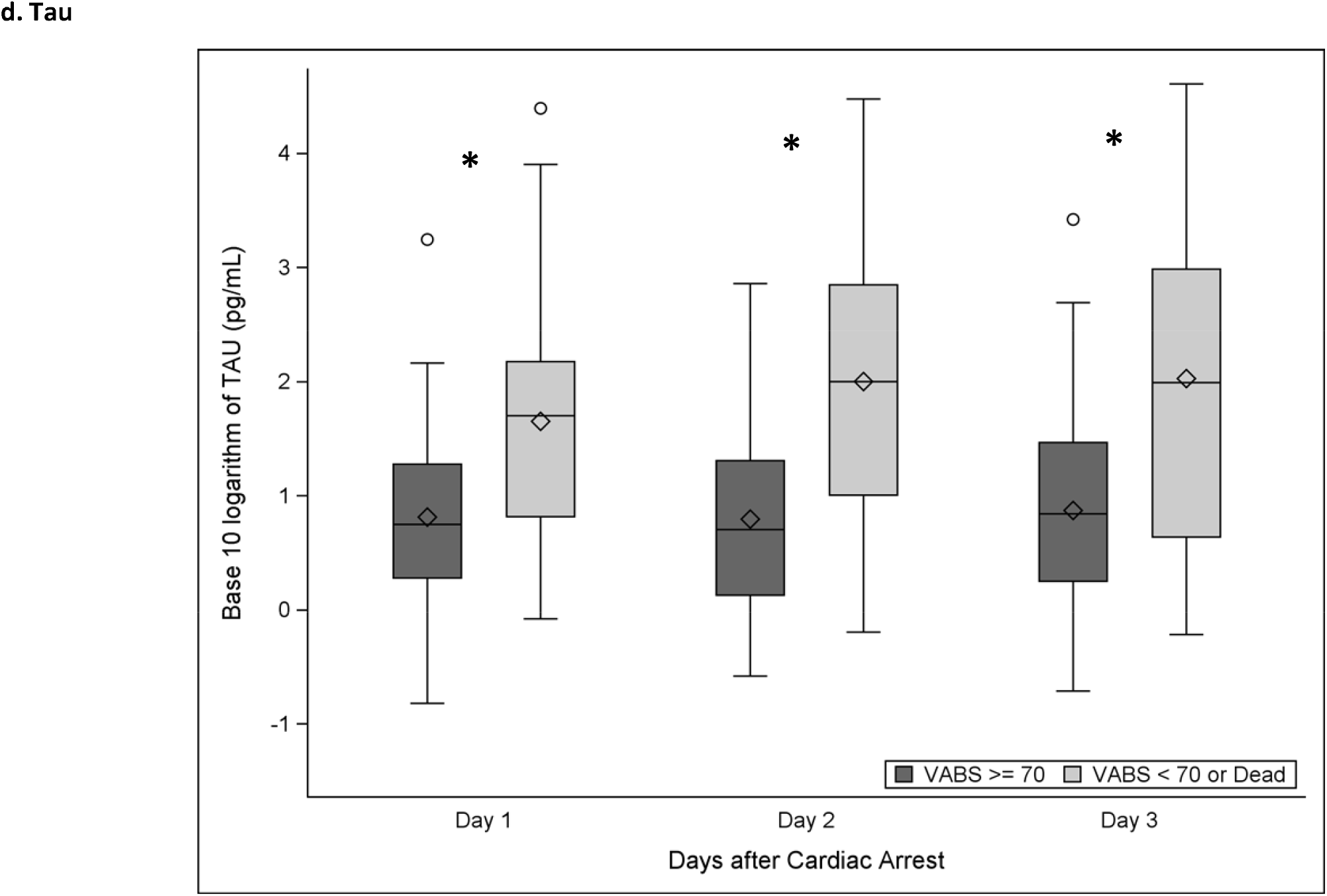
**a-d**. Log transformed biomarker concentrations overall and by outcome group. ^*^p<0.05 for association of biomarker concentration with outcome

### AUROC by outcome group

Figures 3a-c display individual biomarker AUROCs for days 1-3. On day 1, NfL had the highest outcome classification to predict 1 year outcome (AUROC 0.731 [95% confidence interval 0.642, 0.820]), improving to 0.824 (0.742, 0.907) on day 3. UCH-L1 had the highest outcome classification on days 2 (0.860 [0.785, 0.935] and 3 (0.837 [0.747, 0.926]).

**Figure 3.**
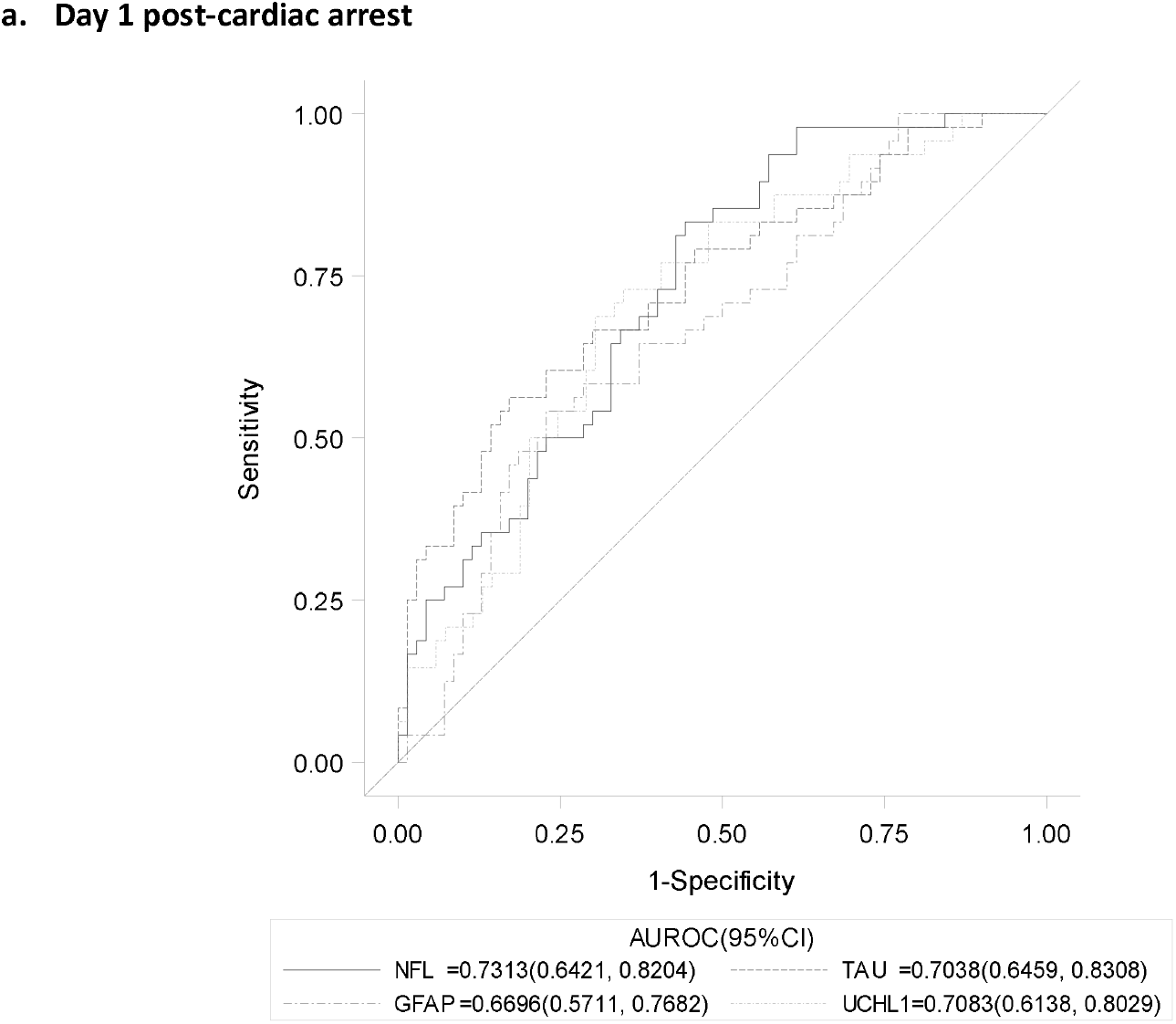
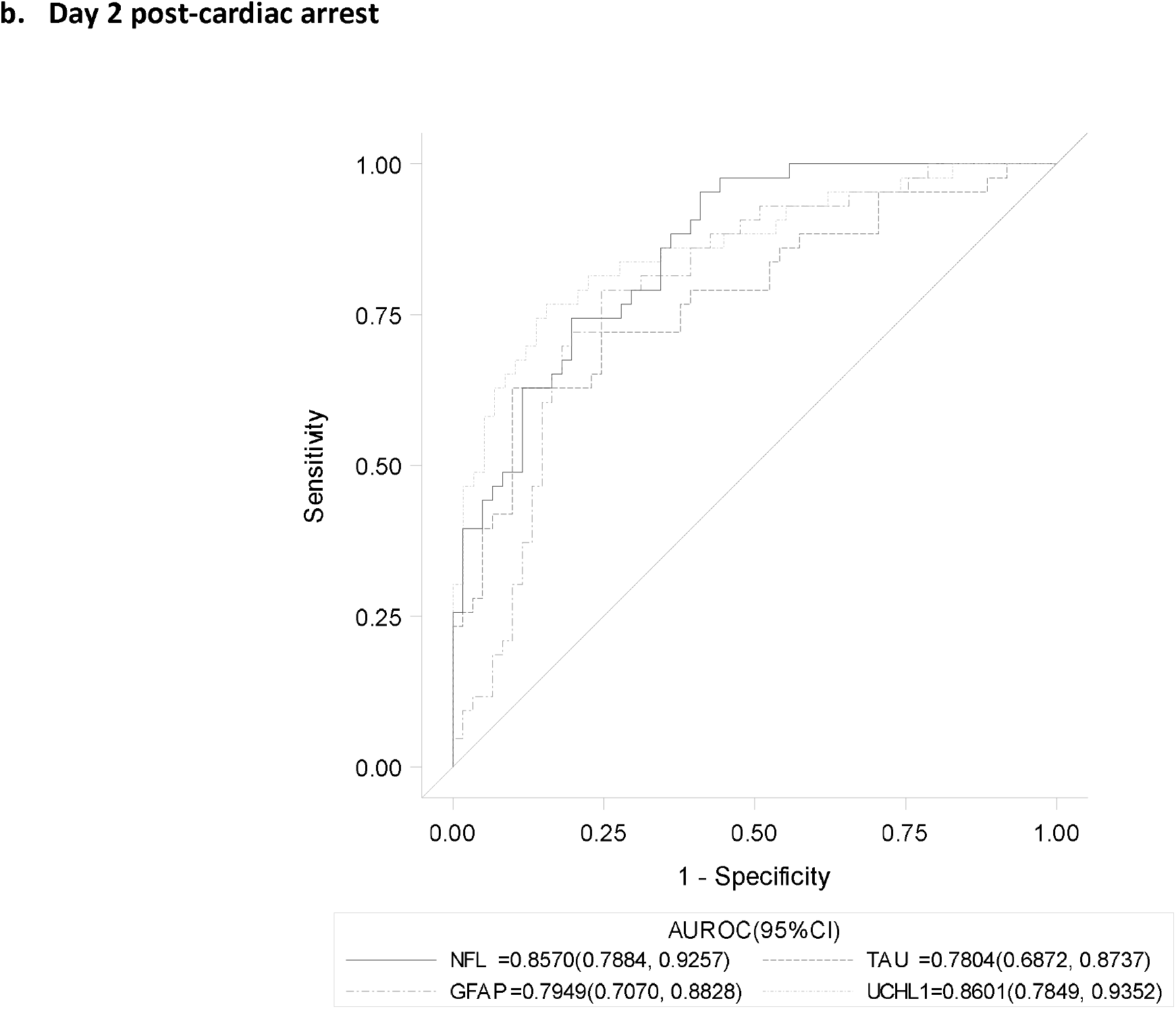
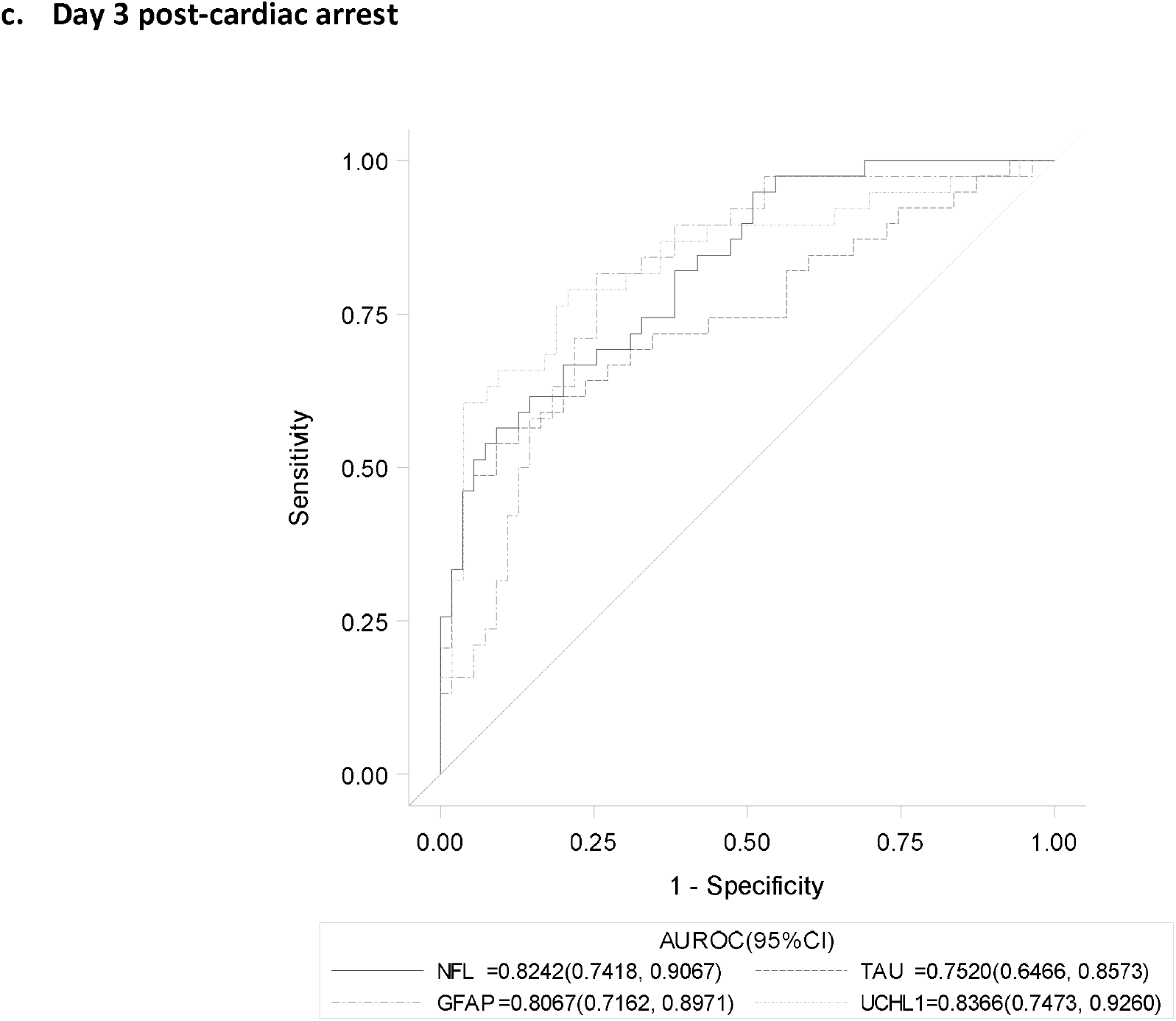
**a-c**. Area under the receiver operator curves for each biomarker on days 1, 2, and 3 and one year unfavorable outcome. GFAP, Glial fibrillary acidic protein; UCH-L1, ubiquitin carboxyl-terminal esterase-L1; NfL, neurofilament light Biomarker concentrations in base 10 log scale.

Biomarker sensitivity and threshold values for each day selected to minimize false positive identification of unfavorable outcomes are reported in Supplemental Table 3. On all study days, 3 children were falsely predicted to have unfavorable outcome by all four biomarkers, except on day 2 where UCH-L1 false positives occurred for 2 children. Tau had the highest sensitivity of the four biomarkers when setting specificity to 95% on day 1 (0.333 [0.204, 0.484]) and UCH-L1 had the best sensitivity on days 2 (0.581 [0.421, 0.730] and 3 (0.605 [0.434, 0.760]).

### Multivariate logistic regressions by biomarker

Multivariate logistic regressions were performed for each day for individual biomarkers to predict one-year outcome with adjustment for patient and clinical covariates (Table 3 [a-d] and Supplemental Table 4 [a-d].). Concentrations of the following were associated with unfavorable one-year outcome: NfL on post-cardiac arrest day 1 (adjusted odds ratio 5.9 [95% confidence interval 1.8, 19.2], day 2 (11.9 [3.8, 36.9]), and day 3 (10.2 [3.1, 33.3], UCH-L1 on day 2 (11.3 [3.0, 42.4]) and day 3 (7.6 [2.1, 27.1]), GFAP on day 2 (2.3 [1.2, 4.5]) and day 3 (2.2 [1.2, 4.0]), and tau on day 1 (2.4 [1.1, 5.3]), day 2 (2.3 [1.3, 4.0]) and day 3 (2.0 [1.2, 3.6]), p<0.05.

**Table 3.** **(a-d)**. Stepwise, multivariate logistic regressions for favorable or unfavorable outcome at 1 year on days 1, 2, and 3 post-cardiac arrest. Entry and stay level of 0.20 and forced inclusion of log biomarker concentrations were used to evaluate the association between individual biomarker concentrations on each day and one-year outcome. ^1^ p < 0.05 for association with outcome on specified study day

|  | Day 1 |  | Day 2 |  | Day 3 |  |
| --- | --- | --- | --- | --- | --- | --- |
|  | Odds Ratio | 95% Wald CI | Odds Ratio | 95% Wald CI | Odds Ratio | 95% Wald CI |
| Neurofilament light, log pg/ml | 5.91 | 1.82, 19.19<sup>1</sup> | 11.88 | 3.82, 36.92<sup>1</sup> | 10.22 | 3.14, 33.33<sup>1</sup> |
| Age, years | 0.90 | 0.79, 1.04 |  |  |  |  |
| Male vs Female sex | 0.35 | 0.10, 1.23<sup>1</sup> | 0.67 | 0.20, 2.31<sup>1</sup> |  |  |
| Cardiac vs Asphyxia etiology | 2.55 | 0.70, 9.34<sup>1</sup> |  |  | 1.96 | 0.50, 7.68<sup>1</sup> |
| Witnessed | 0.15 | 0.03, 0.84<sup>1</sup> |  |  |  |  |
| Pediatric Index of Mortality score | 2.47 | 1.54, 3.97<sup>1</sup> | 2.09 | 1.39, 3.14<sup>1</sup> | 1.92 | 1.29, 2.86<sup>1</sup> |
| TTM used for therapeutic hypothermia | 0.02 | 0.00, 0.76<sup>1</sup> | 0.04 | 0.002, 0.70<sup>1</sup> | 0.08 | 0.004, 1.55 |

b. Ubiquitin carboxyl-terminal esterase- L1
|  | Day 1 |  | Day 2 |  | Day 3 |  |
| --- | --- | --- | --- | --- | --- | --- |
|  | Odds Ratio | 95% Wald CI | Odds Ratio | 95% Wald CI | Odds Ratio | 95% Wald CI |
| Ubiquitin carboxyl-terminal esterase- L1, log pg/ml | 2.01 | 0.84, 4.84 | 11.27 | 3.00, 42.36 | 7.56 | 2.11, 27.09 |
| Age, years | 0.90 | 0.79, 1.01 | 0.91 | 0.79, 1.04 | 0.89 | 0.76, 1.05 |
| Male vs Female sex | 0.30 | 0.09, 1.01 |  |  | 0.56 | 0.14, 2.29 |
| Cardiac vs Asphyxia etiology | 2.61 | 0.76, 8.95 |  |  | 2.00 | 0.47, 8.54 |
| Witnessed | 0.14 | 0.03, 0.81 | 0.09 | 0.01, 0.67 | 0.11 | 0.01, 0.94 |
| Pediatric Index of Mortality score | 2.29 | 1.44, 3.64 | 1.44 | 0.93, 2.21 | 1.56 | 0.97, 2.49 |
| TTM used for therapeutic hypothermia | 0.02 | 0.001, 0.37 | 0.02 | 0.001, 0.62 | 0.02 | 0.001, 0.83 |

c. Glial fibrillary acidic protein
|  | Day 1 |  | Day 2 |  | Day 3 |  |
| --- | --- | --- | --- | --- | --- | --- |
|  | Odds Ratio | 95% Wald CI | Odds Ratio | 95% Wald CI | Odds Ratio | 95% Wald CI |
| Glial fibrillary acidic protein, log pg/ml | 1.36 | 0.72, 2.59 | 2.31 | 1.19, 4.48 | 2.19 | 1.19, 4.03 |
| Age, years | 0.88 | 0.79, 0.99 | 0.90 | 0.79, 1.02 | 0.92 | 0.81, 1.04 |
| Male vs Female sex | 0.30 | 0.09, 0.98 | 0.30 | 0.08, 1.09 | 0.48 | 0.15, 1.60 |
| Cardiac vs Asphyxia etiology | 3.31 | 0.94, 11.65 | 3.82 | 0.94, 15.58 |  |  |
| Witnessed | 0.17 | 0.03, 0.91 | 0.15 | 0.03, 0.87 | 0.13 | 0.02, 0.69 |
| Pediatric Index of Mortality score | 2.47 | 1.56, 3.90 | 1.96 | 1.23, 3.11 | 1.71 | 1.13, 2.58 |
| TTM used for prevention of fever |  |  |  |  | 1.48 | 0.42, 5.18 |
| TTM used for therapeutic hypothermia | 0.02 | 0.001, 0.38 | 2.31 | 1.19, 4.48 | 0.05 | 0.003, 0.92 |

d. Tau
|  | Day 1 |  | Day 2 |  | Day 3 |  |
| --- | --- | --- | --- | --- | --- | --- |
|  | Odds Ratio | 95% Wald CI | Odds Ratio | 95% Wald CI | Odds Ratio | 95% Wald CI |
| Tau, log pg/ml | 2.44 | 1.14, 5.25 | 2.28 | 1.31, 3.97 | 2.04 | 1.16, 3.57 |
| Age, years | 0.90 | 0.80, 1.02 | 0.93 | 0.83, 1.04 | 0.97 | 0.87, 1.10 |
| Male vs Female sex | 0.24 | 0.07, 0.87 | 0.53 | 0.16, 1.73 | 0.58 | 0.19, 1.79 |
| Cardiac vs Asphyxia etiology | 2.25 | 0.65, 7.84 |  |  |  |  |
| Witnessed | 0.11 | 0.02, 0.71 | 0.14 | 0.03, 0.68 |  |  |
| Pediatric Index of Mortality score | 2.10 | 1.35, 3.26 | 1.79 | 1.19, 2.68 | 1.87 | 1.27, 2.75 |
| TTM used for prevention of fever |  |  |  |  | 1.90 | 0.59, 6.14 |
| TTM used for therapeutic hypothermia | 0.02 | 0.001, 0.53 | 0.06 | 0.004, 0.72 | 0.16 | 0.02, 1.17 |

TTM, targeted temperature management; CI, confidence interval
<sup>1</sup> $p < 0.05$ after adjusting for age, sex, race, etiology, location, witnessed, bystander status, ECMO, severity of illness (PIM3), TTM for fever or hypothermia
Multivariate logistic regression for favorable or unfavorable outcomes. Stepwise selection with entry and stay level of 0.20, force the inclusion of log biomarker concentration into models.

Covariates most frequently associated with unfavorable outcomes across the multivariate models were unwitnessed event, higher Pediatric Index of Mortality score, and lack of TTM for hypothermia (Table 3).

## Discussion

In the largest prospective blood-based brain-injury biomarker study conducted in pediatric cardiac arrest to date, each of the four blood-based brain injury biomarkers analyzed early in the post-arrest period discriminated between favorable and unfavorable one-year outcomes with high accuracy^24^. These results remained robust after adjustment for common clinical predictors of outcome including unwitnessed event status and admission risk of mortality score^25^.

NfL and UCH-L1 both demonstrated high overall predictive accuracy (e.g., AUROC) and reliability (e.g., over the time course examined). Concentrations of NfL in both favorable and unfavorable groups numerically increased over the study days, potentially representing ongoing neuroaxonal injury^26, 27^. Our data are consistent with a single center study in children with cardiac arrest with acute respiratory distress syndrome that found NfL predictive of unfavorable outcome at hospital discharge, defined by Pediatric Cerebral Performance Category scores at hospital discharge^15^. A multicenter study that tested biobank samples from a randomized controlled trial in 717 adults with cardiac arrest also found NfL concentrations were the most robust biomarker compared to neuron specific enolase, S100b, and Tau to predict unfavorable outcome as assessed by the Cerebral Performance Category Scale at 6 months^16^. The accuracy of NfL was more robust in adults (AUROC 0.94-0.95 over the first 3 days post-arrest) compared to our study, which could reflect differences related to development, arrest phenotype, as well as patient selection^28^. All patients in the adult trial suffered an out-of-hospital cardiac arrest, had presumed cardiac etiology, and were comatose at the time of recruitment, whereas our study criteria were intentionally broad, with the aim to test biomarker performance across a heterogeneous cohort (e.g., out-of-and in-hospital cardiac arrest, asphyxia and cardiac etiology) of children surviving to ICU admission (without known care limitations) for greater generalizability and to support biomarker translation into general clinical practice.

UCH-L1 and GFAP, both FDA approved for use in mild TBI for clinical decision-making, had highest predictive accuracy on days 2 and 3 post-cardiac arrest, replicating the findings reported in our pilot study^7, 8^. UCH-L1 had the highest sensitivity of the four biomarkers when optimizing specificity on days 2 and 3. Interestingly, median UCH-L1 concentrations remained unchanged over the 3 days in both outcome groups whereas GFAP concentrations increased over the study period in patients with unfavorable outcome. A possible reason for increasing GFAP levels is the fact that it is both released and induced by injured and/or dying astrocytes^23^. Dynamic trajectories may offer more information than a single time point for some biomarkers^29^.

Clinical implications of our study are broad and impactful. First, although most surviving children had favorable 1 year outcome according to the VABS, they remain at risk of cognitive dysfunction^30^. Children may not be neurologically assessable on examination early after resuscitation due to the need for sedative or neuromuscular blockade medications^31^. The primary goal of the POCCA study is to provide clinicians and families with early and accurate tools to assist in prognostication for clinical decision-making. These tools can be used to facilitate a discussion regarding planning for rehabilitative needs or determining goals of care and use of technological support^9^. We found that blood-based brain injury biomarker testing is an accurate method to predict a child’s unfavorable composite outcome of death or poor adaptive behavior functional outcomes at one year. Ours and other groups’ findings could support blood-based biomarker translation into pediatric clinical practice^16^. Additionally, these biomarkers could be used as a tool for predictive enrichment in clinical trials if applied to identify patients most likely (or unlikely) to benefit from a neuroprotective intervention. They may also serve as surrogate outcomes to evaluate responsiveness to interventions^32, 33^. Finally, our future directions include testing blood-based brain injury biomarkers together and with other clinical variables (e.g., unwitnessed event, PIM3 score, TTM) and tests (e.g., brain imaging) to optimize a clinically-relevant prognostication panel for implementation in the acute post-arrest period^34-36^.

### Study limitations

Patient sample size decreased over days 2 and 3 due to deaths and lack of sample availability. Biomarker concentrations were measured in the first 3 days as the study objective was to inform early prognostication; however, concentrations from beyond day 3, alone and together with other biomarkers and clinical variables, may improve prognostication accuracy. Biomarker sampling strategy was pragmatic, acknowledging the need to prevent additional blood draws for safety (e.g., use of leftover laboratory samples and risk of infection to indwelling catheters). Thus, samples occurred within a day’s time rather than at a specific time point post-arrest. Biomarkers were measured by a private company with proprietary assays as clinical laboratory measurements are currently unavailable. Thus, comparisons of biomarker values between this study and others may be difficult to interpret. Detailed neurodevelopmental outcomes (e.g., cognitive) and outcomes past 1 year were not performed, both which could potentially enhance the ability to determine if blood-based brain injury biomarkers are useful to prognosticate specific functional domain impairments. Post-resuscitation care, including the use of TTM, were not standardized in this study. Thus, findings associated with TTM and outcome should be interpreted with caution. Outcome analysis was not adjusted for pre-arrest baseline function. However, children with baseline PCPC 4 or 5 were excluded from the study, which performs similarly to the unfavorable VABS threshold in this study^37^.

## Conclusion

Blood-based brain injury biomarkers prognosticated death or unfavorable adaptive behavior composite outcome at 1 year post-pediatric cardiac arrest with excellent accuracy. Accuracy of biomarkers to predict neurodevelopmental outcomes beyond 1 year should be evaluated.

## Acknowledgements

We acknowledge the **POCCA Investigators**: Karen Hallermeier Walson, MD (Children’s Healthcare of Atlanta); Alexis A Topjian, MD, MSCE (Children’s Hospital of Philadelphia); Christopher Newth, MD (Children’s Hospital of Los Angeles); Elizabeth A Hunt, MD, PhD, Jordan Duval-Arnould, DrPH, MPH (Johns Hopkins Children’s Center); Binod Balakrishnan, MD, Michael T. Meyer, MD, MS, FCCM (Children’s Hospital of Wisconsin); Melissa G Chung, MD (Nationwide Children’s Hospital); Frederick Willyerd, MD (Phoenix Children’s Hospital); Lincoln Smith, MD, Jesse Wenger, MD (Seattle Children’s Hospital); Stuart Friess, MD, Jose Pineda, MD (St. Louis Children’s Hospital); J. Wesley Diddle, MD, Jason Patregnani, MD, Ashley Siems, MD (Children’s National Hospital); Aline B. Maddux, MD, MSCS, Christopher Ruzas, MD, Craig Press, MD, PhD, Peter M. Mourani, MD, (Children’s Hospital of Colorado); Leslie Doughty, MD (Cincinnati Children’s Hospital Medical Center); Juan Piantino, MD, (Oregon Health &Science University); and Ericka L. Fink, MD, MS (UPMC Children’s Hospital of Pittsburgh).

## POCCA Research Coordinators

Beena Desai, BS, CCRC, Maureen G. Richardson, BSN, RN, CPN, Cynthia Bates, CCRP (Children’s Healthcare of Atlanta); Darshana Parikh, Janice Prodell, Maddie Winters, Katherine Smith, MPH, BSN, RN, CPN (Children’s Hospital of Philadelphia); Jeni Kwok, JD, Adriana Cabrales, BA (Children’s Hospital Los Angeles); Ronke Adewale, Pam Melvin(Johns Hopkins Children’s Center); Sadaf Shad, Katherine Siegel, Katherine Murkowski, Mary Kasch (Children’s Wisconsin); Josey Hensley RN, BSN, Lisa Steele, RN, BSN (Nationwide Children’s Hospital); Danielle Brown, Brian Burrows, Lauren Hlivka (Phoenix Children’s Hospital); Deana Rich (Seattle Children’s Hospital); Amila Tutundzic, Tina Day, Lori Barganier (St. Louis Children’s Hospital); Ashley Wolfe, Mackenzie Little, Elyse Tomanio, Neha Patel, Diane Hession (Children’s National Hospital); Yamila Sierra MPH, CCRP (Children’s Hospital of Colorado); Rhonda Jones, Laura Benken (Cincinnati Children’s Hospital Medical Center); and David Maloney, BS, Pamela Rubin, RN (UPMC Children’s Hospital of Pittsburgh).

We also thank Jonathan Elmer, MD, MS; Subramanian Subramanian, MD; Srikala Narayanan, MD; Nicole Toney, MPH, Julia Wallace; Tami Robinson; Andrew Frank; Stefan Bluml, PhD; Jessica Wisnowski, PhD; Keri Feldman; Avinash Vemulapalli; Linda Ryan; Scott Szypulski, MBA; Christopher Keys, and all of the children and families who generously participated in POCCA. We are grateful to all the ICU and other hospital staff, nurses, and physicians for their efforts in study recruitment and provision of outstanding clinical care to families.

## Supplemental Materials

Online only Tables 1-4

## Figure Legends

**Figure 1.** Enrollment Diagram.

**Figure 2 a-d**. Biomarker concentrations by overall and outcome group.

**Figure 3 a-c**. Area under the receiver operator curves for each biomarker on days 1, 2, and 3.

