## Supplemental tables 1-4 for "Blood-Based Brain Injury Biomarkers to Prognosticate Outcome after Pediatric Cardiac Arrest"

**SUPPLEMENTAL MATERIAL**

**Supplemental table 1.** Post-resuscitation hospital data overall and by favorable and unfavorable outcome groups. ICU, intensive care unit; CPR, cardiopulmonary resuscitation. p-value are based on Chi-square test for categorical variables, Kruskal-Wallis test for continuous variables. Favorable outcome was Vineland Adaptive Behavioral Scale-III (VABS) score >=70 and Unfavorable outcome was VABS < 70 or died.

| N (%) or Median (interquartile range) | Overall N=120 | Favorable N=70 | Unfavorable N=50 | p-value |
| --- | --- | --- | --- | --- |
| **Fever (> 38°C)** | 59 (49.2) | 34 (48.6) | 25 (50.0) | 0.877 |
| **Seizure occurrence** | 33 (27.5) | 12 (17.1) | 21 (42.0) | 0.003 |
| **Arterial blood gas values, day 1** |  |  |  |  |
| Lowest PaO2, mmHg | 63.5 (48.0, 83.0)  N=94 | 60.0 (46.5, 74.8)  N=52 | 71.0 (55.0, 88.0)  N=42 | 0.062 |
| Highest PaO2, mmHg | 193.5 (105.0, 283.7)  N=94 | 235.5 (119.0, 325.0)  N=52 | 169.8 (100.0, 239.0)  N=42 | 0.048 |
| Lowest PaCO2, mmHg | 30.4 (27.0, 36.0)  N=94 | 30.0 (27.7, 35.9)  N=52 | 30.7 (26.0, 36.5)  ]n=42 | 0.948 |
| Highest PaCO2, mmHg | 50.2 (42.5, 63.7)  N=94 | 53.5 (43.5, 67.5)  N=52 | 44.0 (38.7, 52.0)  N=50 | 0.003 |
| **Lowest systolic blood pressure, mmHg** | 67.0 (55.5, 79.5)  N=120 | 67.5 (58.0, 80.0)  N=70 | 65.5 (52.0, 78.0)  N=50 | 0.377 |
| **Highest systolic blood pressure, mmHg** | 124.0 (111.0, 141.5)  N=120 | 122.0 (109.0, 140.0)  N=70 | 125.5 (114.0, 144.0)  N=50 | 0.583 |
| **Invasive mechanical ventilation** | 117 (97.5) | 67 (95.7) | 50 (100.0) | 0.138 |
| **Invasive mechanical ventilation, days** | 8.0 (4.0, 16.0)  N=117 | 8.0 (4.0, 13.0)  N=67 | 9.5 (4.0, 22.0)  N=50 | 0.278 |
| **Vasoactive infusions, day 1** |  |  |  |  |
| Epinephrine | 76 (63.33) | 76 (63.33) | 76 (63.33) | 0.370 |
| Milrinone | 28 (23.33) | 17 (24.29) | 11 (22.00) | 0.770 |
| Norepinephrine | 22 (18.33) | 10 (14.29) | 12 (24.00) | 0.175 |
| Dopamine | 13 (10.83) | 5 (7.14) | 8 (16.00) | 0.124 |
| Vasopressin | 12 (10.00) | 5 (7.14) | 7 (14.00) | 0.217 |
| Phenylephrine | 6 (5.00) | 4 (5.71) | 2 (4.00) | 0.671 |
| Isoproterenol | 1 (0.83) | 1 (1.43) | 0 (0) | 0.396 |
| Dobutamine | 0 | 0 | 0 | NA |
| **First lactate, mmol/L** | 5.8 (2.9, 10.9)  N=107 | 4.8 (2.3, 8.9)  N=62 | 8.6 (3.8, 13.2)  N=45 | 0.014 |
| **Pediatric Index of Mortality, %** | 16 (7-31) | 14 (5-18) | 29 (15-88) | <0.001 |
| **First Glasgow Coma Scale score in the ICU** | 4 (3, 9)  N=88 | 7 (3, 11)  N=48 | 3 (3, 7)  N=40 | 0.016 |
| **Extracorporeal membrane oxygenation** | 28 (23.3) | 18 (25.7) | 10 (20.0) | 0.466 |
| Extracorporeal CPR | 22 (78.6) | 15 (83.3) | 7 (70.0) | 0.410 |
| **Target temperature management, prevention of fever** | 31 (25.8) | 12 (17.1) | 18 (36.0) | 0.032 |
| **Target temperature management, therapeutic hypothermia** | 15 (12.5) | 12 (17.1) | 3 (6.0) | 0.069 |
| Duration at target temperature, h | 59 (40, 82) | 59 (41, 82) | 62 (19, 130) | 0.942 |

**Supplemental Table 2 (a-c).** Unadjusted biomarker concentrations on days 1-3 overall and by outcome group. All biomarker concentrations are in pg/ml. GFAP, Glial fibrillary acidic protein; UCH-L1, ubiquitin carboxyl-terminal esterase- L1; NfL, neurofilament light

a. **Day 1**

| Median (interquartile range) | **Overall N=120** | **Favorable** **N=70** | **Unfavorable** **N=50** | **Kruskal-Wallis**  **p-value** |
| --- | --- | --- | --- | --- |
| **NFL** | n=118 | n=70 | n=48 | <0.001 |
|  | 27.95 (9.88, 70.75) | 13.81 (6.29, 49.98) | 50.54 (20.56, 169.00) |  |
| **Tau** | n=118 | n=70 | n=48 | <0.001 |
|  | 8.97 (2.85, 60.70) | 5.59 (1.89, 18.85) | 50.10 (6.53, 149.30) |  |
| **GFAP** | n=118 | n=70 | n=48 | 0.002 |
|  | 212.51 (109.00, 816.76) | 174.85 (79.63, 401.27) | 469.88 (137.70, 2780.48) |  |
| **UCH-L1** | n=118 | n=70 | n=48 | <0.001 |
|  | 138.95 (40.04, 518.77) | 73.39 (25.36, 230.77) | 310.40 (110.73, 848.36) |  |

b. **Day 2**

| Median (interquartile range) | **Overall N=120** | **Favorable** **N=70** | **Unfavorable** **N=50** | **Kruskal-Wallis**  **p-value** |
| --- | --- | --- | --- | --- |
| **NFL** | n=104 | n=61 | n=43 | <0.001 |
|  | 75.45 (20.08, 299.66) | 31.17 (13.15, 88.67) | 287.26 (78.23, 898.47) |  |
| **Tau** | n=104 | n=61 | n=43 | <0.001 |
|  | 15.17 (2.40, 108.41) | 5.04 (1.34, 20.23) | 99.59 (10.09, 702.00) |  |
| **GFAP** | n=104 | n=61 | n=43 | <0.001 |
|  | 342.01 (139.43, 1912.20) | 162.39 (104.46, 361.89) | 1515.27 (386.16, 16470.00) |  |
| **UCH-L1** | n=104 | n=61 | n=43 | <0.001 |
|  | 94.18 (32.80, 285.75) | 51.31 (22.85, 98.31) | 319.10 (163.24, 2129.17) |  |

c. **Day 3**

| Median (interquartile range) | **Overall N=120** | **Favorable** **N=70** | **Unfavorable** **N=50** | **Kruskal-Wallis**  **p-value** |
| --- | --- | --- | --- | --- |
| **NFL** | n=94 | n=55 | n=39 | <0.001 |
|  | 177.60 (35.33, 469.38) | 60.63 (23.59, 217.99) | 490.88 (136.52, 1512.99) |  |
| **Tau** | n=94 | n=55 | n=39 | <0.001 |
|  | 15.44 (2.63, 103.01) | 6.92 (1.78, 29.16) | 97.65 (4.33, 964.00) |  |
| **GFAP** | n=93 | n=55 | n=38 | <0.001 |
|  | 334.30 (150.55, 5205.00) | 166.21 (98.36, 404.55) | 2736.71 (368.75, 12812.05) |  |
| **UCH-L1** | n=93 | n=55 | n=38 | <0.001 |
|  | 117.72 (32.75, 344.54) | 49.67 (21.43, 133.10) | 357.36 (188.96, 1737.00) |  |

**Supplemental Table 3 (a-c).** Biomarker specificity, threshold level (pg/ml), sensitivity on univariate biomarker models, with set specificity of 95%. Biomarkers on base-10 logarithm scale.

a. **Day 1**

| **Model** | **Specificity (95%CI)** | **Threshold level** | | **Sensitivity (95%CI)** | **Patients, N (%)** | | | | |
| --- | --- | --- | --- | --- | --- | --- | --- | --- | --- |
|  |  | (pg/mL) | Base 10 logarithm |  | True Positive | False Negative | False Positive | True Negative | Total |
| **NFL** | 0.957 (0.880, 0.991) | 184.00 | 2.26 | 0.250 (0.136, 0.396) | 12 (10.17) | 36 (30.51) | 3 (2.54) | 67 (56.78) | 118 |
| **Tau** | 0.957 (0.880, 0.991) | 111.75 | 2.05 | 0.333 (0.204, 0.484 | 16 (13.56) | 32 (27.12) | 3 (2.54) | 67 (56.78) | 118 |
| **GFAP** | 0.957 (0.880, 0.991) | 129544.00 | 5.11 | 0.043 (0.005, 0.143) | 2 (1.69) | 46 (38.98) | 3 (2.54) | 67 (56.78) | 118 |
| **UCH-L1** | 0.957 (0.878, 0.991) | 2319.00 | 3.37 | 0.146 (0.061, 0.278) | 7 (5.98) | 41 (35.04) | 3 (2.56) | 66 (56.41) | 117 |

b. **Day 2**

| **Model** | **Specificity (95%CI)** | **Threshold level** | | **Sensitivity (95%CI)** | **Patients, N (%)** | | | | |
| --- | --- | --- | --- | --- | --- | --- | --- | --- | --- |
|  |  | (pg/mL) | Base 10 logarithm |  | True Positive | False Negative | False Positive | True Negative | Total |
| **NFL** | 0.951 (0.863, 0.990) | 389.01 | 2.59 | 0.442 (0.291, 0.601) | 19 (18.27) | 24 (23.08) | 3 (2.88) | 58 (55.77) | 104 |
| **Tau** | 0.951 (0.863, 0.990) | 240.15 | 2.38 | 0.395 (0.259, 0.556) | 17 (16.35) | 26 (25.00) | 3 (2.88) | 58 (55.77) | 104 |
| **GFAP** | 0.951 (0.863, 0.990) | 92690.00 | 4.97 | 0.12 (0.04, 0.25) | 5 (4.81) | 38 (36.54) | 3 (2.88) | 58 (55.77) | 104 |
| **UCH-L1** | 0.948 (0.856, 0.989) | 276.15 | 2.44 | 0.581 (0.421, 0.730) | 25 (24.75) | 18 (17.82) | 3 (2.97) | 55 (54.46) | 101 |

c. **Day 3**

| **Model** | **Specificity (95%CI)** | **Threshold level** | | **Sensitivity (95%CI)** | **Patients, N (%)** | | | | |
| --- | --- | --- | --- | --- | --- | --- | --- | --- | --- |
|  |  | (pg/mL) | Base 10 logarithm |  | True Positive | False Negative | False Positive | True Negative | Total |
| **NFL** | 0.946 (0.849, 0.989) | 490.88 | 2.69 | 0.513 (0.348, 0.676) | 20 (21.28) | 19 (20.21) | 3 (3.19) | 52 (55.32) | 94 |
| **Tau** | 0.946 (0.849, 0.989) | 108.48 | 2.04 | 0.487 (0.324, 0.652) | 19 (20.21) | 20 (21.28) | 3 (3.19) | 52 (55.32) | 94 |
| **GFAP** | 0.946 (0.849, 0.989) | 40040.00 | 4.60 | 0.211 (0.096, 0.373) | 8 (8.60) | 30 (32.26) | 3 (3.23) | 52 (55.91) | 93 |
| **UCH-L1** | 0.962 (0.870, 0.995) | 341.60 | 2.53 | 0.605 (0.434, 0.760) | 23 (25.27) | 15 (16.48) | 2 (2.20) | 51 (56.04) | 91 |

**Supplemental Table 4 (a-d)**. Univariate logistic regressions for favorable or unfavorable outcome at 1 year by blood-based brain injury biomarker on post-cardiac arrest day 1, 2, and 3. TTM, targeted temperature management; CI, confidence interval; NA, not applicable

a. **Neurofilament light**

|  | **Day 1** | | **Day 2** | | **Day 3** | |
| --- | --- | --- | --- | --- | --- | --- |
|  | **Odds**  **Ratio** | **95% Wald CI** | **Odds**  **Ratio** | **95% Wald CI** | **Odds**  **Ratio** | **95% Wald CI** |
| Neurofilament light, log pg/ml | 4.23 | 2.12, 8.45 | 11.55 | 4.52, 29.54 | 7.76 | 3.22, 18.69 |
| Age, years | 0.98 | 0.92, 1.04 | 0.98 | 0.92, 1.04 | 0.98 | 0.92, 1.04 |
| Male vs Female sex | 0.92 | 0.44, 1.93 | 0.92 | 0.44, 1.93 | 0.92 | 0.44, 1.93 |
| Non-White vs White Race | 0.82 | 0.38, 1.79 | 0.82 | 0.38, 1.79 | 0.82 | 0.38, 1.79 |
| Cardiac vs Asphyxia etiology | 0.89 | 0.38, 2.07 | 0.89 | 0.38, 2.07 | 0.89 | 0.38, 2.07 |
| In-hospital vs Out of hospital event | 0.24 | 0.11, 0.52 | 0.24 | 0.11, 0.52 | 0.24 | 0.11, 0.52 |
| Witnessed | 0.09 | 0.03, 0.26 | 0.09 | 0.03, 0.26 | 0.09 | 0.03, 0.26 |
| Bystander resuscitation | 3.38 | 1.54, 7.41 | 3.38 | 1.54, 7.41 | 3.38 | 1.54, 7.41 |
| Extracorporeal membrane oxygenation | 0.72 | 0.30, 1.73 | 0.72 | 0.30, 1.73 | 0.72 | 0.30, 1.73 |
| Pediatric Index of Mortality score | 2.10 | 1.53, 2.89 | 2.10 | 1.53, 2.89 | 2.10 | 1.53, 2.89 |
| TTM used for prevention of fever | 2.47 | 1.07, 5.68 | 2.47 | 1.07, 5.68 | 2.47 | 1.07, 5.68 |
| TTM used for therapeutic hypothermia | 0.31 | 0.08, 1.16 | 0.31 | 0.08, 1.16 | 0.31 | 0.08, 1.16 |

b. **Ubiquitin carboxyl-terminal esterase- L1**

|  | **Day 1** | | **Day 2** | | **Day 3** | |
| --- | --- | --- | --- | --- | --- | --- |
|  | **Odds**  **Ratio** | **95% Wald CI** | **Odds**  **Ratio** | **95% Wald CI** | **Odds**  **Ratio** | **95% Wald CI** |
| Neurofilament light, log pg/ml | 2.85 | 1.61, 5.05 | 16.32 | 5.13, 51.92 | 7.14 | 2.90, 17.60 |
| Age, years | 0.98 | 0.92, 1.04 | 0.98 | 0.92, 1.04 | 0.98 | 0.92, 1.04 |
| Male vs Female sex | 0.92 | 0.44, 1.93 | 0.92 | 0.44, 1.93 | 0.92 | 0.44, 1.93 |
| Non-White vs White Race | 0.82 | 0.38, 1.79 | 0.82 | 0.38, 1.79 | 0.82 | 0.38, 1.79 |
| Cardiac vs Asphyxia etiology | 0.89 | 0.38, 2.07 | 0.89 | 0.38, 2.07 | 0.89 | 0.38, 2.07 |
| In-hospital vs Out of hospital event | 0.24 | 0.11, 0.52 | 0.24 | 0.11, 0.52 | 0.24 | 0.11, 0.52 |
| Witnessed | 0.09 | 0.03, 0.26 | 0.09 | 0.03, 0.26 | 0.09 | 0.03, 0.26 |
| Bystander resuscitation | 3.38 | 1.54, 7.41 | 3.38 | 1.54, 7.41 | 3.38 | 1.54, 7.41 |
| Extracorporeal membrane oxygenation | 0.72 | 0.30, 1.73 | 0.72 | 0.30, 1.73 | 0.72 | 0.30, 1.73 |
| Pediatric Index of Mortality score | 2.10 | 1.53, 2.89 | 2.10 | 1.53, 2.89 | 2.10 | 1.53, 2.89 |
| TTM used for prevention of fever | 2.47 | 1.07, 5.68 | 2.47 | 1.07, 5.68 | 2.47 | 1.07, 5.68 |
| TTM used for therapeutic hypothermia | 0.31 | 0.08, 1.16 | 0.31 | 0.08, 1.16 | 0.31 | 0.08, 1.16 |

c. **Glial fibrillary acidic protein**

|  | **Day 1** | | **Day 2** | | **Day 3** | |
| --- | --- | --- | --- | --- | --- | --- |
|  | **Odds**  **Ratio** | **95% Wald CI** | **Odds**  **Ratio** | **95% Wald CI** | **Odds**  **Ratio** | **95% Wald CI** |
| Neurofilament light, log pg/ml | 1.69 | 1.11, 2.57 | 2.57 | 1.60, 4.14 | 2.74 | 1.66, 4.50 |
| Age, years | 0.98 | 0.92, 1.04 | 0.98 | 0.92, 1.04 | 0.98 | 0.92, 1.04 |
| Male vs Female sex | 0.92 | 0.44, 1.93 | 0.92 | 0.44, 1.93 | 0.92 | 0.44, 1.93 |
| Non-White vs White Race | 0.82 | 0.38, 1.79 | 0.82 | 0.38, 1.79 | 0.82 | 0.38, 1.79 |
| Cardiac vs Asphyxia etiology | 0.89 | 0.38, 2.07 | 0.89 | 0.38, 2.07 | 0.89 | 0.38, 2.07 |
| In-hospital vs Out of hospital event | 0.24 | 0.11, 0.52 | 0.24 | 0.11, 0.52 | 0.24 | 0.11, 0.52 |
| Witnessed | 0.09 | 0.03, 0.26 | 0.09 | 0.03, 0.26 | 0.09 | 0.03, 0.26 |
| Bystander resuscitation | 3.38 | 1.54, 7.41 | 3.38 | 1.54, 7.41 | 3.38 | 1.54, 7.41 |
| Extracorporeal membrane oxygenation | 0.72 | 0.30, 1.73 | 0.72 | 0.30, 1.73 | 0.72 | 0.30, 1.73 |
| Pediatric Index of Mortality score | 2.10 | 1.53, 2.89 | 2.10 | 1.53, 2.89 | 2.10 | 1.53, 2.89 |
| TTM used for prevention of fever | 2.47 | 1.07, 5.68 | 2.47 | 1.07, 5.68 | 2.47 | 1.07, 5.68 |
| TTM used for therapeutic hypothermia | 0.31 | 0.08, 1.16 | 0.31 | 0.08, 1.16 | 0.31 | 0.08, 1.16 |

d. **Tau**

|  | **Day 1** | | **Day 2** | | **Day 3** | |
| --- | --- | --- | --- | --- | --- | --- |
|  | **Odds**  **Ratio** | **95% Wald CI** | **Odds**  **Ratio** | **95% Wald CI** | **Odds**  **Ratio** | **95% Wald CI** |
| Neurofilament light, log pg/ml | 2.97 | 1.78, 4.96 | 3.03 | 1.88, 4.90 | 2.59 | 1.63, 4.12 |
| Age, years | 0.98 | 0.92, 1.04 | 0.98 | 0.92, 1.04 | 0.98 | 0.92, 1.04 |
| Male vs Female sex | 0.92 | 0.44, 1.93 | 0.92 | 0.44, 1.93 | 0.92 | 0.44, 1.93 |
| Non-White vs White Race | 0.82 | 0.38, 1.79 | 0.82 | 0.38, 1.79 | 0.82 | 0.38, 1.79 |
| Cardiac vs Asphyxia etiology | 0.89 | 0.38, 2.0) | 0.89 | 0.38, 2.07 | 0.89 | 0.38, 2.07 |
| In-hospital vs Out of hospital event | 0.24 | 0.11, 0.52 | 0.24 | 0.11, 0.52 | 0.24 | 0.11, 0.52 |
| Witnessed | 0.09 | 0.03, 0.26 | 0.09 | 0.03, 0.26 | 0.09 | 0.03, 0.26 |
| Bystander resuscitation | 3.38 | 1.54, 7.41 | 3.38 | 1.54, 7.41 | 3.38 | 1.54, 7.41 |
| Extracorporeal membrane oxygenation | 0.72 | 0.30, 1.73 | 0.72 | 0.30, 1.73 | 0.72 | 0.30, 1.73 |
| Pediatric Index of Mortality score | 2.10 | 1.53, 2.89 | 2.10 | 1.53, 2.89 | 2.10 | 1.53, 2.89 |
| TTM used for prevention of fever | 2.47 | 1.07, 5.68 | 2.47 | 1.07, 5.68 | 2.47 | 1.07, 5.68 |
| TTM used for therapeutic hypothermia | 0.31 | 0.08, 1.16 | 0.31 | 0.08, 1.16 | 0.31 | 0.08, 1.16 |
